# Radiomics Analysis for Predicting Growth of Subsolid Lung Nodules on CT

**DOI:** 10.1101/2023.12.05.23299456

**Authors:** Shiny Weng, Masha Bondarenko, Gunvant Chaudhari, Arun Innaje, Terrence Chen, Brandon K.K. Fields, Jae Ho Sohn

**Affiliations:** University of California, Berkeley; Department of Radiology and Biomedical Imaging, University of California, San Francisco; United Imaging Intelligence, Boston

## Abstract

**Background:** Accurate identification of growing subsolid nodules is crucial for effective risk stratification and the early detection of invasive lung cancer, allowing for timely treatment while avoiding unnecessary surgery on low-risk nodules that would otherwise remain stable. The traditional method of risk stratification, which relies on qualitative visual analysis of CT scans, remains challenging. Therefore, this study aims to leverage a longitudinal dataset of subsolid nodules on CT and develop radiomic and clinical feature-based models to identify nodules that are likely to grow over time.

**Purpose:** The purpose of this study is to develop a machine learning model to predict the growth of subsolid nodules using a combination of radiomic and clinical features.

**Materials and Methods:** A retrospective study was conducted on a cohort of patients who had undergone chest CT scans at a single institution between 2015 and 2019. Corresponding radiology reports were used to extract ground truth labels for the nodules’ growth status (i.e., increased, decreased, unchanged), as well as other related information such as nodule size, nodule type, location, slice location, component sizes, and compared studies. An automated NLP pipeline was employed for the extraction of labels. Additionally, associated CT scans were processed through a commercial nodule characterization algorithm, which generated parameters such as nodule size, location, and 3D segmentation coordinates. Utilizing this dataset, along with radiomic features computed from pyRadiomics and clinical features (e.g., patient age and sex), models for predicting the growth of subsolid nodules were developed. The primary metric used to evaluate model performance was the AUC-ROC, assessed on both the independent validation set and averaged across 5-fold cross-validation. A total of 15 features were used for the model, selected through a combination of recursive feature elimination, random forest importance analysis, and univariate selection during cross-validation.

**Results:** The final dataset comprises 950 subsolid nodules, each with corresponding growth statuses and 3D segmentations (799 unchanged, 139 growing, 12 decreasing). Among various models, support vector machine (SVM) achieved the highest AUC of 0.81 on both 5-fold cross-validation and the independent validation set. Furthermore, in the statistical analysis of the association between radiomic and clinical features and increasing versus unchanged nodules, 88 radiomic features were identified as statistically significant (p < 0.05) through a Mann-Whitney U test, with 82 of these radiomic features being highly statistically significant (p < 0.01). Notable radiomic features, including Voxel Volume, Run Length Non-Uniformity, and Dependence Non-Uniformity, demonstrated high feature importance in identifying growing nodules in the SVM model. Among the clinical features, Patient Age exhibited high feature importance.

**Conclusion:** A model based on combined radiomic and clinical features was trained to predict subsolid nodules that would grow over time. Dependence Non-Uniformity, Run Length Non-Uniformity, Voxel Volume, Gray Level Non-Uniformity, and Patient Age were among the most predictive features for identifying high-risk, growing nodules.

## Introduction

Lung cancer remains a significant global health concern and consistently ranks among the top causes of cancer-related mortality [1]. Early or pre-neoplasia in the form of subsolid nodules on CT poses a clinical conundrum for the thoracic tumor board due to their heterogeneous risk of invasion, metastasis, and growth over time. Predicting the growth of lung nodules is of high clinical importance because it enables more precise targeted treatment of high-risk nodules at an early stage while allowing low-risk nodules to be either left untouched or longitudinally followed over time [2]. The traditional method of subsolid nodule risk stratification involves qualitative visual analysis of nodules over months to years through repeated CT scans by radiologists. However, this method has multiple disadvantages, including but not limited to the risk of patient loss to follow-up, an overly extended time for characterization, and causing patient anxiety during the follow-up period [3]–[5]. The importance of early detection in achieving positive patient outcomes, as well as the increase in subsolid nodule identification in CT, further emphasizes the need to address this challenge [4].

Ensuring the precise evaluation and timely treatment of growing subsolid nodules will significantly contribute to the survival of individuals with lung cancer. Consequently, given the urgency of these challenges, radiomics, situated at the intersection of medical imaging, data science, and advanced image analysis, has garnered substantial attention as a quantitative approach to nodule characterization [6]. This innovative approach entails the extraction of a comprehensive array of quantitative features from both 3D and 2D medical images, potentially revealing complex patterns hidden from the naked eye. Moreover, as evidenced by previous studies, radiomics has been successfully employed to address various clinical problems, including the classification between benign and malignant nodules [7]–[10]. In a study of 86 part-solid ground glass nodules, Chae *et al*. successfully differentiated invasive lung adenocarcinoma from preinvasive lesions by employing a combination of texture-based features [11]. In Ma *et al.*, the authors identified 583 features of nodule intensity, heterogeneity, and shape, which they employed to achieve 82.7% accuracy in the classification of malignant and benign pulmonary nodules [12].

In the realm of radiomic studies, widely used multivariate models, such as linear and quadratic discriminant analysis, logistic regression, support vector machines, and random forests classifiers, have been frequently employed [6], [8], [9], [13]. These models, when combined with clinical and/or demographic features, transform radiomics into a powerful tool capable of uncovering intricate characteristics within pulmonary nodule images [13], [14]. Modern machine learning techniques can leverage this wealth of information to develop accurate prediction models, marking a significant stride in the comprehensive understanding and effective management of lung cancer.

Our objective is to develop a machine learning model that leverages both clinical and radiomic features extracted from chest CT scans to accurately identify growing adenocarcinoma spectrum nodules, thereby assisting the thoracic tumor board in optimizing their management and improving overall outcomes.

## Materials and Methods

### Dataset

In this retrospective and Institutional Review Board (IRB) approved study, chest CT scans were collected from a cohort of patients who had undergone imaging at a single institution between 2015 and 2019. The range of CT scans encompassed low-dose, non-contrast, contrast-enhanced, and high-resolution chest CTs, as well as CT angiograms obtained from various sites and different vendors. Corresponding radiology reports were utilized to extract ground truth labels for the nodules’ growth status (i.e., increased, decreased, unchanged), along with other pertinent information such as nodule size, nodule type (e.g., pure ground glass vs. mixed solid/ground glass), location (e.g., lobe of the lung), slice location, component sizes, and compared studies.

A total of 1737 consecutive and comprehensively collected radiology reports describing subsolid nodules were initially retrieved using multiple keyword-based searches. An automated NLP pipeline, employing regular expression and heuristics, was utilized for label extraction, and manual review was conducted to ensure the accuracy of the dataset.

To link the nodules reported in the radiology reports with their specific coordinates and segmentations, we processed the CT scans using a commercial algorithm (United Imaging Intelligence, Boston, MA, USA), which identified the location, coordinates, and nodule segmentations. We then applied a matching algorithm using the parameters generated by the commercial algorithm and those extracted from the radiology reports to identify the intersection of the two groups. A workflow pipeline is displayed in *Figure 2*.

A subsolid nodule’s ‘growth’ was defined as follows: 1) increased total size, 2) new or growing solid component, or 3) increasing density of the nodule. Additionally, a board-certified radiologist and a second-year radiology resident manually verified the set of growing nodules.

### Radiomics

All images were converted to the NIfTI format for use in pyRadiomics. Using the results of the commercial algorithm and its intersection with the radiology report dataset, an image processing algorithm was applied to locate the nodule within the CT scan. A 3D bounding box with dimensions (50 px, 50 px, nodule thickness) was generated, centered around the nodule and volume of interest. From these bounding boxes, the 3D bounded image and its segmentation mask were extracted.

Following this approach, pyRadiomics (v3.1.0), an open-source package for extracting features from 2D and 3D medical images [15], was employed to generate first-order, shape, and texture-based radiomic features for each nodule. This process of creating the image and mask allowed the extraction of relevant radiomic features and characteristics for subsequent analysis. The list of radiomic features can be found in pyRadiomics’ documentation, and sample information derived from commercial algorithms and report text is presented in *Table 2*.

### Data Processing and Preparation

Due to the incomplete and inconsistent nature of free-text radiology reports and the false-positive identification of acute ground glass nodular opacities by the commercial algorithm, a graduated intersection algorithm was applied to identify those that were most confidently matched on a scale from 1 to 12. Out of the 4025 nodules identified in the radiology reports, 950 nodules from the report text were confidently extracted. These nodules were confirmed to be true subsolid nodules as described in the radiology reports, correctly segmented by the commercial algorithm, and validated as a match between the report and the algorithm.

To ensure greater accuracy and confidence in a nodule’s reported growth status, we applied a 180-day rule: nodules reported as unchanged from a previous scan that occurred less than or equal to 180 days ago were removed from the dataset before training [16].

### Feature Analysis, Engineering, and Selection

To enhance model performance, various feature engineering techniques were applied. These encompassed one-hot encoding for categorical variables (e.g., patient sex), feature scaling, and standardization, as well as polynomial and logarithmic transformations. Moreover, various data visualizations, such as correlation heatmaps, feature importance plots, and box plots, were utilized in combination with domain knowledge to account for collinearities and correlations in the data [6]. Additionally, a filtering algorithm based on statistical methods, such as Mann-Whitney U tests,

Chi-squared tests, and pairwise correlation analyses with a threshold of 0.8 (Pearson correlation), was employed, and recursive feature elimination was utilized to generate a set of relevant features. In the analysis of radiomic and clinical features, various univariate tests were conducted, including non-parametric Mann-Whitney U tests using Python’s SciPy 1.8.1 ‘mannwhitneyu’ function. These tests aimed to analyze and quantify the differences between the increasing and unchanging nodule groups.

### Model Training

As observed in many medical datasets, a significant class imbalance dilemma was noted within the dataset. Specifically, in the nodule dataset assembled for this study, approximately 15% (139) of nodules were classified as increasing. Therefore, to split the dataset into training, testing, and validation sets, the ‘train_test_split’ function from scikit-learn was used, which has the ability to stratify based on the output label. To address the significant class imbalance, 5-fold cross-validation was employed to verify and analyze the model’s results [17]. Furthermore, hyperparameter optimization was conducted using scikit-learn’s ‘GridSearchCV.’ Various models recommended in prior studies, including support vector machine, adaptive boosting (AdaBoost), random forests, logistic regression, and residual neural network, were explored for training [6], [9]. SVM was chosen as the optimal model, and all algorithms were programmed using Python (version 3.8), scikit-learn (version 1.3), and PyTorch (version 2.0).

### Model Evaluation

A total of 15 features were selected for the model using a combination of recursive feature elimination, random forest importance analysis, and univariate selection during cross-validation. The primary metric for evaluating model performance was the AUC-ROC, assessed on both the independent validation set and averaged across 5-fold cross-validation. The selected threshold for the SVM classifier was chosen by optimizing for the F1 score, and a confusion matrix was analyzed to assess the tradeoff between precision and recall, offering a more comprehensive evaluation for clinical decision-making [18].

### Error Analysis

Error analysis was conducted by assessing the tradeoff between precision and recall in a confusion matrix. Additionally, a Mann-Whitney U test was applied to assess differences between the true-positive and false-negative groups, providing statistical insights into potential distinctions in their distributions. To complement these quantitative analyses, a qualitative visual analysis is presented in *Figure 3*.

## Results

### Dataset

The intersection dataset generated from radiology reports and the commercial algorithm consists of 1234 subsolid nodules, with ground truth labels (i.e., increase, unchanged, decrease) available for 950 of them. There are 799 (∼84%) unchanged nodules, 139 (∼15%) increasing nodules, and 12 (∼1%) decreasing nodules. A detailed breakdown of the dataset and corresponding patient characteristics is presented in *Table 1*.

**Table 1.**

| Dataset and Patient Characteristics |  |  |  |
| --- | --- | --- | --- |
| Characteristics | Unchanging Group<br>(N = 799, 85.2%) | Increasing Group<br>(N = 139, 14.8%) | P-Value § |
| Age — year (% > 60) *† | 70.3 ± 10.6 (84.3%) | 73.1 ± 11.6 (86.3%) | <0.001 |
| Male Sex — no. (%) | 239 | 47 | 0.411 |
| Nodule Size — mm. ‡ | 6.9 ± 4.0 | 10.4 ± 4.6 | <0.001 |
| Nodule Volume — mm<sup>3</sup> | 440.3 ± 1085.3 | 1072.0 ± 1353.0 | <0.001 |

\* Plus-minus values are means ± SD † The percentage is the proportion of patients with age > 60 out of both unchanging and increasing groups, respectively ‡ Nodule Size is the average diameter of the nodule from its long and short axes § P-value is computed from Mann-Whitney U tests (increasing vs. unchanging) and Chi-squared tests (male vs. female)

### Feature Analyses

Results from non-parametric Mann-Whitney U tests indicated that 88 radiomic features were statistically significant (p < 0.05), with 82 of them being highly statistically significant (p < 0.01). Notably, features that demonstrated significant importance included Voxel Volume and Gray Level Non-Uniformity, as illustrated in *Table 2*. As validated in this study and seen in prior studies, nodule size remains a widely and highly predictive indicator for assessing the probability of nodule malignancy [28]. Additionally, a Chi-squared test was conducted to evaluate Patient Sex (∼70% Female vs. ∼30% Male), but ultimately, this analysis did not yield statistically significant differences between nodule statuses.

**Table 2.**

| SVM Features |  |  |  |  |
| --- | --- | --- | --- | --- |
| # | Feature Name | Feature Type | Radiomic Family | P-Value † |
| 1 | Patient Age | Clinical | N/A | <0.001 |
| 2 | Maximum 2D Diameter Row | Radiomic | Shape | <0.001 |
| 3 | Maximum 3D Diameter | Radiomic | Shape | <0.001 |
| 4 | Maximum 2D Diameter Slice | Radiomic | Shape | <0.001 |
| 5 | Maximum 2D Diameter Column | Radiomic | Shape | <0.001 |
| 6 | Surface Volume Ratio | Radiomic | Shape | <0.001 |
| 7 | Surface Area | Radiomic | Shape | <0.001 |
| 8 | Voxel Volume | Radiomic | Shape | <0.001 |
| 9 | Zone Entropy | Radiomic | GLSZM | <0.001 |
| 10 | Gray Level Non-Uniformity | Radiomic | GLSZM | <0.001 |
| 11 | Correlation | Radiomic | GLCM | <0.001 |
| 12 | Inverse Difference Normalized | Radiomic | GLCM | <0.001 |
| 13 | Run Length Non-Uniformity | Radiomic | GLRLM | <0.001 |
| 14 | Dependence Entropy | Radiomic | GLDM | <0.001 |
| 15 | Dependence Non-Uniformity | Radiomic | GLDM | <0.001 |

\* The above features were used in the SVM model, resulting in an AUC of 0.81 on both independent validation and 5-fold cross-validation.
† The p-value is computed by performing a Mann-Whitney U test between two groups: increasing nodules and unchanging nodules.

### Model Evaluation

We implemented various machine learning models, with SVM (utilizing a radial basis function kernel) emerging as the top-performing model, achieving an AUC of 0.81 on both the 5-fold cross-validation and the independent validation set. To assist physicians and readers in better interpreting AUC scores, a confusion matrix is presented in *Figure 1*. The confusion matrix was computed using the top-performing cross-validated model on the independent validation set. This model was tuned with a False Positive Rate (FPR) threshold of 0.23, which was chosen by optimizing for the F1 score. Nearly all progressive nodules in the independent validation set were correctly identified. Furthermore, as observed in the statistical analyses and exploratory data analysis of the radiomic features, and as noted in prior research studies [19], many of these features were highly correlated. Consequently, after performing feature dimensionality reduction, the SVM model performed best using only 15 features, as demonstrated in *Figure 1*.

**Figure 1.**
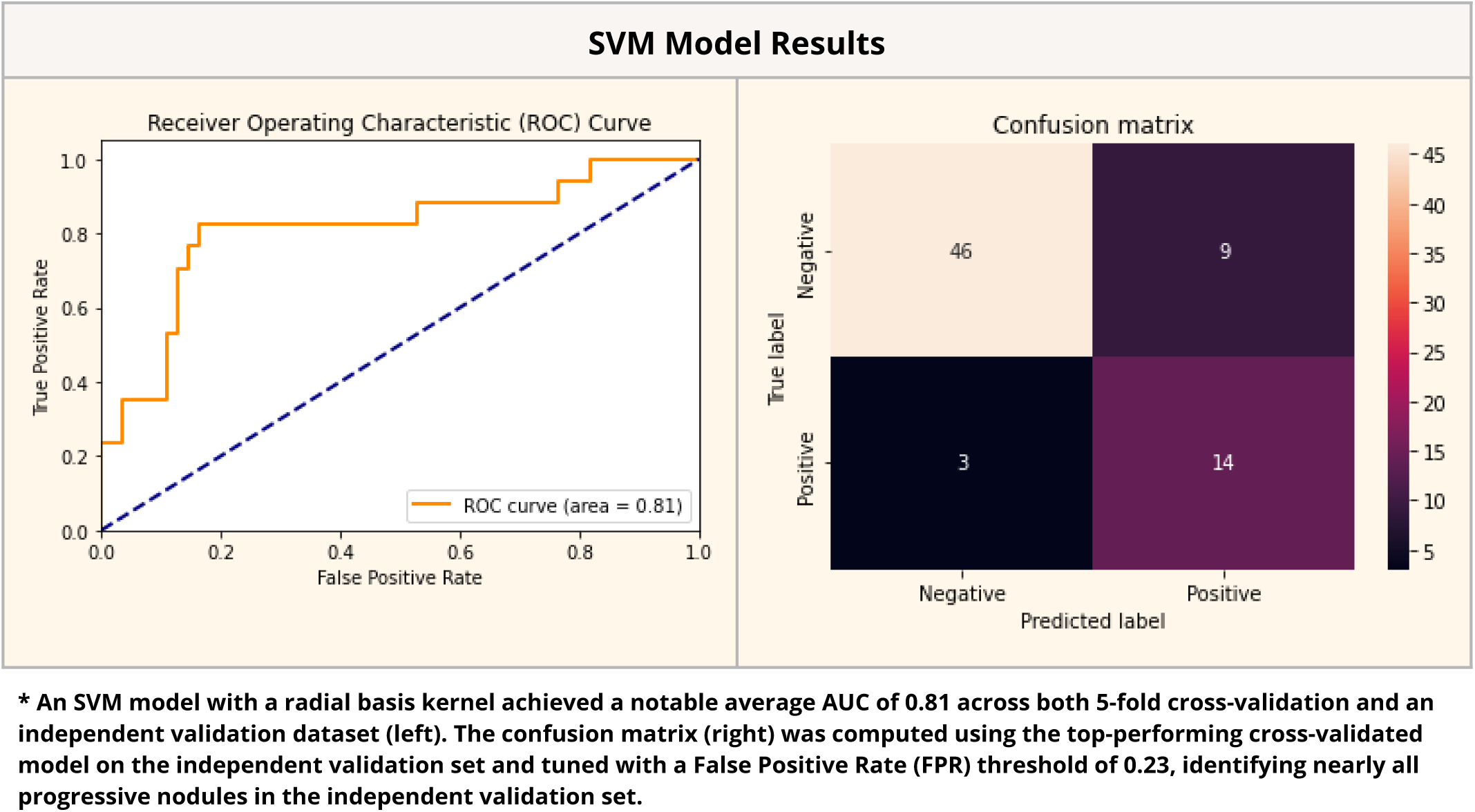

### Error Analysis

The primary focus for this model was to identify true-positive cases of nodule growth, due to its high significance in clinical decision-making. As shown in *Figure 1*’s confusion matrix, instances of false positives and false negatives occurred. Out of 17 positive cases in the independent validation set, 14 were correctly classified as growing nodules, while 3 were incorrectly classified as stable nodules. A Mann-Whitney U test between the true-positive and false-negative cases indicated high statistical significance for each feature used in the SVM model. *Figure 3* displays a gallery of incorrectly classified nodules.

**Figure 2.**
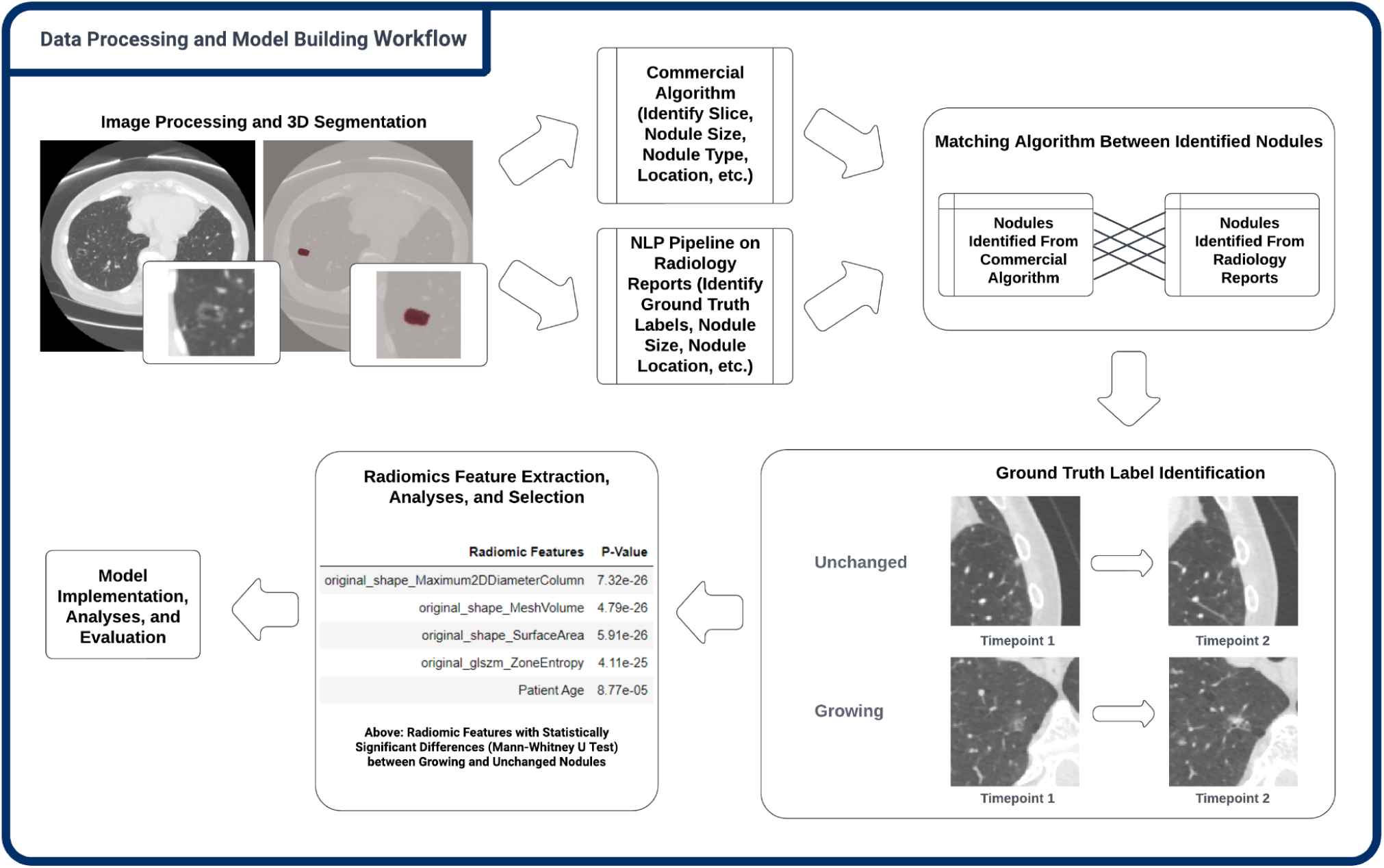

**Figure 3.**
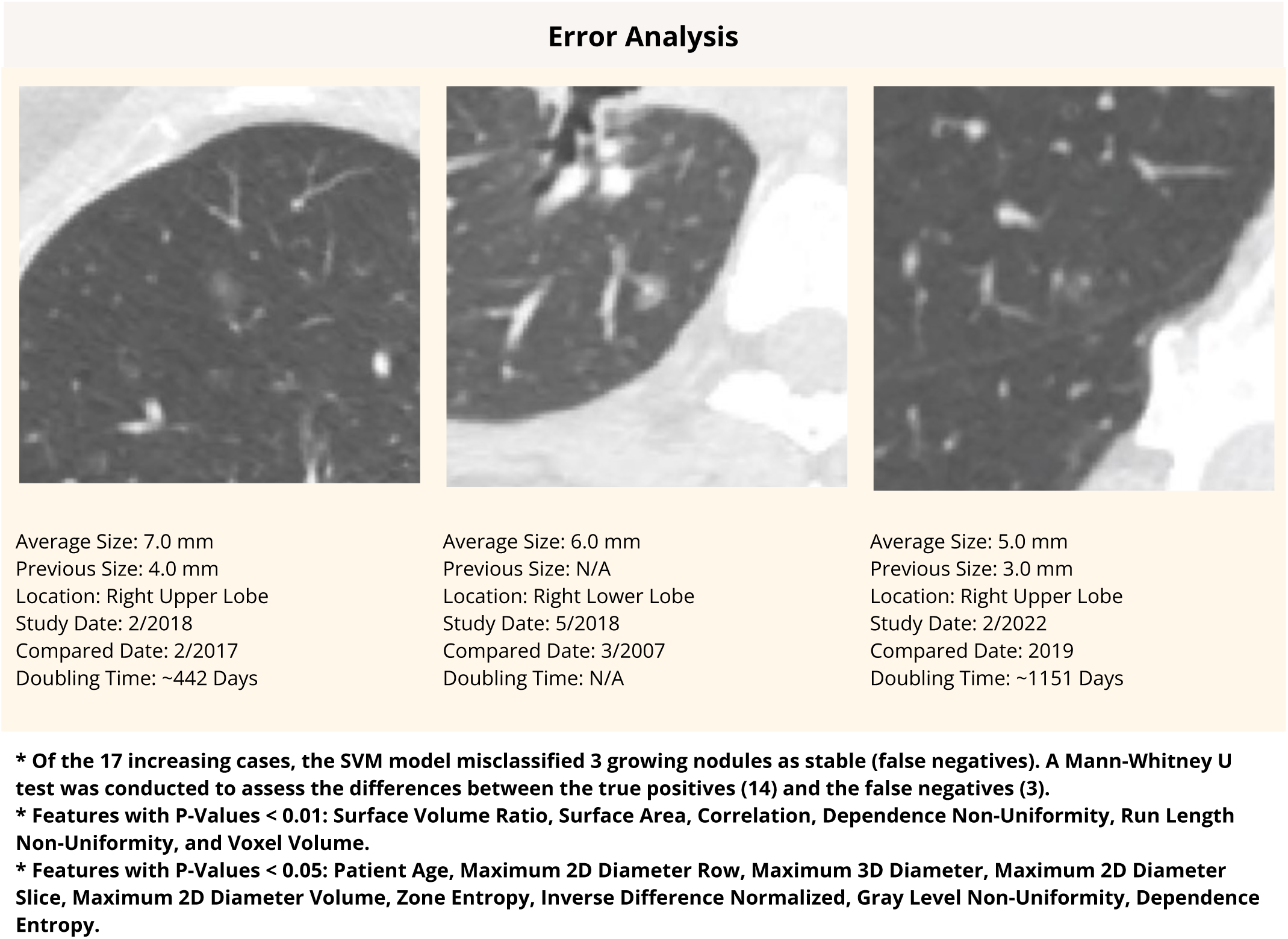

**Figure 4.**
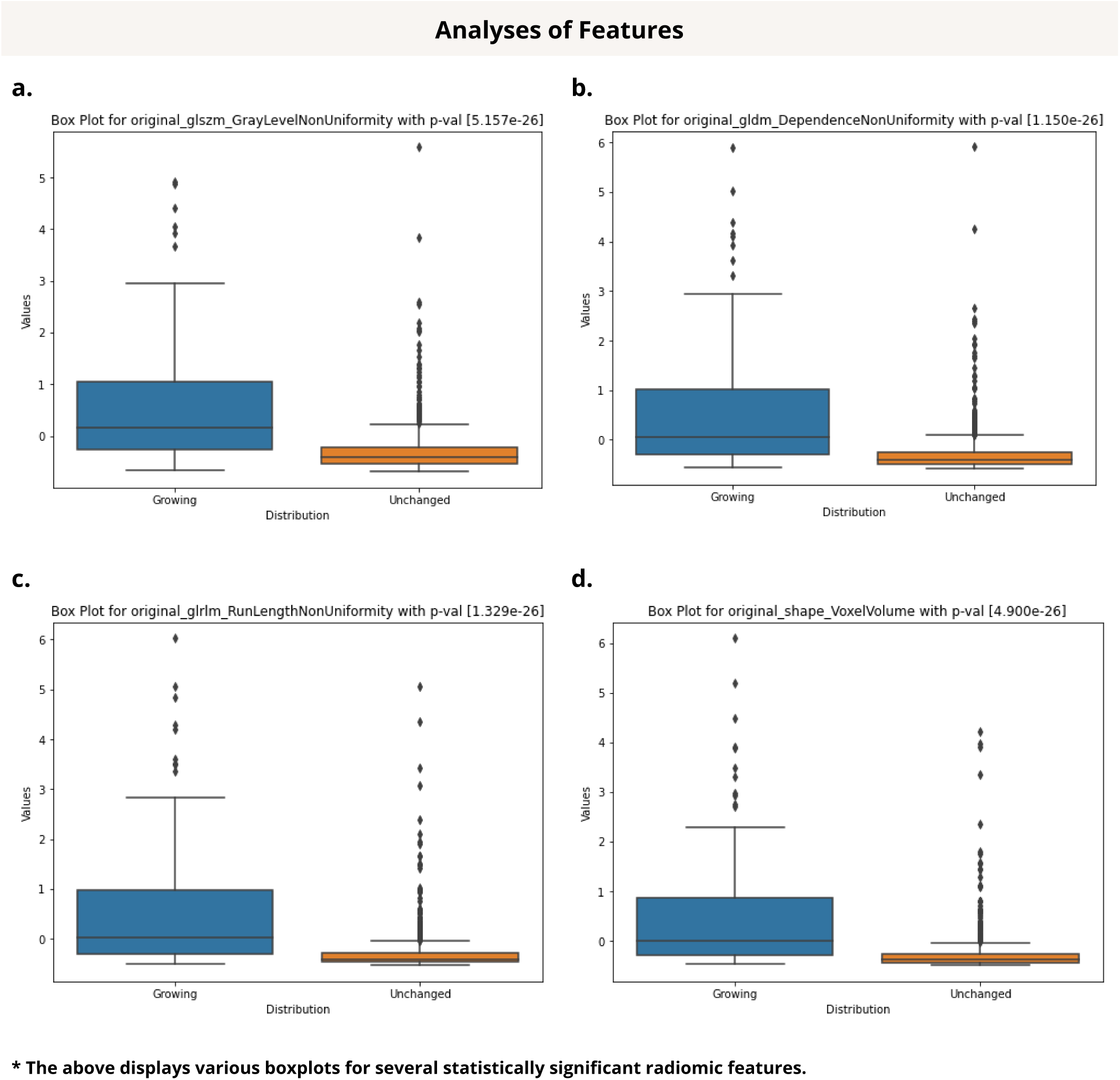

## Discussion

This study underscores the practical importance of integrating radiomic and clinical attributes to predict subsolid nodule growth in CT scans. When managing early or pre-neoplasia of the lungs, especially in the context of deciding whether surgical intervention is necessary, no definitive and widely accepted guidelines exist to inform these management decisions. Such decisions ultimately reside on the availability of local surgical and/or alternative expertise, as well as the practice culture of the individual institution. Although radiological guidelines, such as the Fleischner Society criteria for incidental lung nodules and Lung-RADS for lung cancer screening, provide some guidance on the next steps in management, they do not conclusively determine these critical treatment decisions [16], [20].

The decision to operate or opt for conservative management of pulmonary nodules typically depends on both clinical and imaging factors, including total nodule size, solid component size, and the overall cardiopulmonary health of the patient. The past growth and potential future growth of these nodules over time become increasingly relevant factors in the decision-making process. While the passage of time would naturally serve as the most accurate assessment of nodule growth, this evaluation only occurs once the nodule has already grown. Hence, the interplay between radiomic features, which delve into the intricate characteristics of nodules using advanced imaging analysis, and clinical attributes like patient history and risk factors, can offer a more comprehensive and nuanced understanding of nodule behavior and growth. By combining these two sources of information and leveraging a large dataset of 950 nodules, the developed model has the potential to enhance radiologists’ ability to predict which nodules are likely to grow over time. This, in turn, can provide more accurate and personalized recommendations for the thoracic tumor board and patients.

Compared to existing studies in the literature, our study utilized a large curated dataset of 950 subsolid nodules, all of which were longitudinally confirmed, 3D segmented, and manually verified, although not all pathology confirmed. However, arguably, this dataset is subject to much less selection bias than the cases that are surgically resected and pathology confirmed. This is because most surgically resected datasets would include nodules that were radiologically and clinically deemed highly suspicious, with patients healthy enough to undergo surgery. Moreover, nodules pathologically compatible with adenocarcinoma spectrum lesions do not necessarily indicate growth and invasion in patients, whereas longitudinal observation would arguably represent another potentially more informative way of assessing the risk status of a nodule. Continued surveillance and analysis of pulmonary nodules over time constitutes a critical aspect of nodule management, as underscored by established guidelines and widely adopted practices in clinical settings. The emphasis on longitudinal observation stems from the recognition that the evolution of nodules is a dynamic process, influenced by various factors such as growth patterns, changes in morphology, and alterations in radiographic characteristics [3], [5]. Tao *et al.* predicted nodule invasiveness by utilizing a longitudinal dataset of lung nodules, arguing that results should consider the rich longitudinal data that clinicians acquire during follow-up to identify and track patterns that may otherwise remain uncaptured [21]. Similarly, Paez *et al.* argue for the potential of longitudinal modeling to enhance radiomic predictions of lung cancer by leveraging distinct trends in nodule morphology and size over time [22].

Our most effective model, employing an SVM with a radial basis kernel, achieved an impressive average AUC of 0.81 in both 5-fold cross-validation and independent validation. To underscore the model’s effectiveness, it correctly identified 14 out of 17 growing nodules in the independent validation dataset. The model utilizes the radiomic and clinical features at a single time point to predict growth. Additionally, when assessing the visual interpretation of radiomic features, the set of radiomic features that performed the best collectively hinted at patterns potentially discernible to the naked eye, such as a higher level of surface heterogeneity in progressive and possibly malignant nodules. Various statistical analyses, including Mann-Whitney U tests and correlation analyses, demonstrated the high statistical significance of radiomic features in distinguishing between growing and stable nodules.

To delve deeper into the interpretation of these features, it was observed that, on average, patients with growing nodules were found to be older than those with stable nodules, aligning with the widely recognized notion that age is considered a crucial risk factor in many health contexts [13]. One of the radiomic features examined was Run Length Non-Uniformity (RLN), which exhibited an approximately 336% increase for growing nodules compared to stable nodules on average. RLN measures the dissimilarity in intensity among consecutive pixels or elements in an image, with lower RLN values indicating that the same color or intensity persists over a relatively long distance without interruption. Similarly, Zone Entropy (ZE), a feature that quantifies the uncertainty and randomness in the distribution of zone sizes and gray levels, was found to be approximately 528% higher for growing nodules than for stable nodules on average, indicating a greater degree of heterogeneity in the texture patterns of growing nodules. Other features, such as Gray Level

Non-Uniformity, Inverse Difference Normalized, and Voxel Volume, also provided valuable insights. Collectively, these features suggest that progressive nodules may often appear less homogeneous and more heterogeneous to the naked eye, frequently exhibiting larger sizes [23]. Understanding these patterns could provide practitioners with valuable quantitative insights for determining the suspiciousness of a nodule.

Multiple studies have attempted to use qualitative imaging features to predict growth, but none has been widely accepted, primarily due to low to moderate accuracy at best, which may also stem from a lack of clear qualitative image-based signal and/or small sample size. Our results are consistent with previous studies [7], [13], [14], but are distinctive due to a significantly larger dataset, robust predictive power, utilization of a combination of radiomic and clinical features, and automated data collection methodologies. Our findings underscore the effective performance of our model in both the training and validation sets.

While our study achieved success, it also unveiled several limitations inherent in the nature of radiology reports. Specifically, these limitations encompass discrepancies in reporting, metrics that are either unquantifiable or missing, and subtle uncertainties within radiology report text, including the presence of words such as “may,” “possibly,” and “slightly.” The ground truth label generation process was based solely on the reporting radiologists’ assessment within the report text. Thus, these factors have the potential to influence the consistency of the ground truth labels assigned to nodule growth statuses. Consequently, this underscores the need for improved standards in radiology reporting [24]. High interreader variability poses significant challenges in achieving consistent and accurate diagnoses of pulmonary nodules [25], [26]. Additionally, this highlights the promising implications of combining NLP and computer vision pipelines to validate radiologists’ reportings, provide additional insights, and potentially uncover undetected findings.

It is also worth noting the challenging aspects of curating large datasets for machine learning and deep-learning-based algorithms, especially in medical contexts. The requirement for large volumes of data is further driven by the high complexity and variability of medical data, as well as the diverse range of patient cases and conditions, making it especially challenging to collect, particularly in specialized or rare medical scenarios [27]. Factors such as the limited availability of radiologists and the laborious annotation processes further contribute to the complexity of acquiring suffcient data. Other obstacles include privacy and ethical concerns related to the sensitive patient information involved, the heterogeneity of data from various sources, class imbalance issues, and the need to adhere to strict regulatory compliance. However, by leveraging AI-powered tools, such as automated report generation, image interpretation, and workflow optimization, we may mitigate such issues in data collection.

In conclusion, this study developed a predictive model for identifying the overall nodule growth status, leveraging a combination of radiomic and clinical features. The model achieved an AUC of 0.81 in both cross-validation and an independent validation set. Additionally, we curated a valuable dataset comprising 1234 subsolid nodules, complete with segmentations, ground truth labels, and various other attributes (e.g., nodule size, type, location, slice location) for 950 nodules.

## Author Contributions

Conceptualization: S.W., J.H.S.; Methodology: S.W, J.H.S, B.F.; Data Curation: S.W, M.B., A.I., T.C.; Data Validation: S.W, M.B., J.H.S, B.F.; Data Analysis: S.W.; Resources: S.W, J.H.S, B.K.K.F.; Code and Visualization: S.W, G.R.C.; Manuscript Review/Editing: S.W, J.H.S., B.K.K.F.

## Abbreviations

NLP: (Natural Language Processing)
SVM: (Support Vector Machine)
GLCM: (Gray Level Co-Occurrence Matrix)
GLRLM: (Gray Level Run Length Matrix)
GLSZM: (Gray Level Size Zone Matrix)
GLDM: (Gray Level Dependence Matrix)
(NGTDM): Neighbouring Gray Tone Difference Matrix
ROC: (Receiver Operating Characteristic)
AUC-ROC: (Area Under the ROC Curve)
(NIfTI): Neuroimaging Informatics Technology Initiative

## Data Availability

Data produced by this study contains confidential information about patients and is not publicly available.

## Notes

### Competing Interest Statement

Research collaboration with United Imaging Intelligence.

### Funding Statement

This study did not receive any funding.

### Author Declarations

The Institutional Review Board of University of California, San Francisco gave ethical approval for this work

### Summary of Updates

Author name updated

